# Association of results of four lateral flow antibody tests with subsequent SARS-CoV-2 infection

**DOI:** 10.1101/2022.05.19.22275126

**Authors:** Lucy Findlater, Adam Trickey, Hayley E Jones, Amy Trindall, Sian Taylor-Phillips, Ranya Mulchandani, EDSAB-HOME investigators, Isabel Oliver, David Wyllie

## Abstract

**Background:** SARS-CoV-2 vaccine coverage remains incomplete, being only 15% in low income countries. Rapid point of care tests predicting SARS-CoV-2 infection susceptibility in the unvaccinated might assist in risk management and vaccine prioritisation.

**Methods:** We conducted a prospective cohort study in 2,826 participants working in hospitals and Fire and Police services in England, UK, during the pandemic (ISRCTN5660922). Plasma taken at recruitment in June 2020 was tested using four lateral flow immunoassay (LFIA) devices and two laboratory immunoassays detecting antibodies against SARS-CoV-2 (UK Rapid Test Consortium’s AbC-19™ Rapid Test, OrientGene COVID IgG/IgM Rapid Test Cassette, SureScreen COVID-19 Rapid Test Cassette, and Biomerica COVID-19 IgG/IgM Rapid Test; Roche N and EUROIMMUN S laboratory assays). We monitored participants for microbiologically-confirmed SARS-CoV-2 infection for 200 days. We estimated associations between test results at baseline and subsequent infection, using Poisson regression models adjusted for baseline demographic risk factors for SARS-CoV-2 exposure.

**Findings:** Positive IgG results on each of the four LFIAs were associated with lower rates of subsequent infection: adjusted incidence rate ratios (aIRRs) 0.00 (95% confidence interval 0.00-0.01), 0.03 (0.02-0.05), 0.07 (0.05-0.10), and 0.09 (0.07-0.12) respectively. The protective association was strongest for AbC-19 and SureScreen. The aIRR for the laboratory Roche N antibody assay at the manufacturer-recommended threshold was similar to those of the two best performing LFIAs at 0.03 (0.01-0.10).

**Interpretation:** Lateral flow devices measuring SARS-CoV-2 IgG predicted disease risk in unvaccinated individuals over 200 day follow-up. The association of some LFIAs with subsequent infection was similar to laboratory immunoassays.

**Funding:** UK Government

**Research in context:** *Evidence before this study:* We searched PubMed for research articles, using the search terms (“COVID-19” OR “SARS-CoV-2” OR “2019-nCoV” OR “coronavirus”) AND (“Antibody” OR “IgG”) AND ((“protection” OR “infection”) identifying studies of cohorts of unvaccinated individuals which reported antibody-associated disease protection published between Dec 1 2019 and 1 April 2022. Additionally, we reviewed studies matching “SARS-CoV-2” and “lateral flow” and “antibody” over the same period. Multiple cohort studies in healthy populations have demonstrated an association between the detection of antibodies to SARS-CoV-2 following natural infection and protection from subsequent symptomatic infection with SARS-CoV-2. Protection estimates were about 85% protection in two overlapping meta-analyses, while in several larger studies increased protection with higher antibody levels was observed. Lateral flow immunoassays (LFIAs) detecting anti-SARS-CoV-2 IgG are a cheap, readily deployed technology which has been used on a large scale in population screening programs. However, there are wide variations in sensitivity and specificity of antibody detection between different devices. No studies have investigated whether LFIA results are associated with subsequent SARS-CoV-2 infection.

*Added value of this study:* In a prospective cohort study of 2,826 UK key workers, we found positivity in lateral flow test results had a strong negative association with subsequent SARS-CoV-2 infection within 200 days in an unvaccinated population. The performance of different devices in predicting disease protection differed: positivity on more specific but less sensitive tests was associated with markedly decreased rate of disease. By contrast, protection associated with testing positive using more sensitive devices detecting lower levels of anti-SARS-CoV-2 IgG was more modest.

*Implications of all the available evidence:* If the field performance of these tests against contemporary SARS-CoV-2 infection was similar to that observed in this study, lateral flow tests with high specificity may have a role in estimation of SARS-CoV-2 disease risk in unvaccinated populations and individuals.

## Introduction

The COVID-19 pandemic has caused a global health crisis. Worldwide, as of 14 April 2022, there have been over 490 million cases and 6 million deaths associated with SARS-CoV-2(1). The infection is common worldwide and effective vaccines have now been developed and distributed, most widely in high income countries(2). However, while about 65% of the world’s population have now received at least one vaccine dose, in low income countries the corresponding figure is 15.2%(3). Additionally, vaccine hesitancy is widespread globally(4, 5). Understanding an individual’s risk impacts both hesitancy(4) and other aspects of individual behaviour. Understanding individual risk also contributes to population surveillance, the prioritisation of vaccine delivery, and to pandemic planning and response(5-7).

Antibodies to the SARS-CoV-2 spike and nucleocapsid proteins are generated in over 90% of individuals with symptomatic infection and persist for months(8-13). Multiple studies following up individuals with SARS-CoV-2 infection have described protection to SARS-CoV-2 following natural infection in individuals with detectable antibody levels: protection from subsequent symptomatic infection with SARS-CoV-2 was estimated at about 85% protection in two overlapping meta-analyses of 19 studies performed in the general population, healthcare workers, college students, and residents in long term care facilities(14, 15). In some studies, quantitative antibody levels were recorded, and increased protection in individuals with higher antibody levels was observed(9, 16).

A range of laboratory-based immunoassays and lateral flow immunoassays (LFIAs) have been developed to detect anti-SARS-CoV-2 IgG or IgM antibodies(17-19). LFIAs are small devices that allow antibody testing without the need for a laboratory(18, 20-22), and have been deployed at population scale(23). Previous work has explored the sensitivity and specificity of LFIAs to detect antibody responses to the spike or nucleoprotein antigens of SARS-CoV-2 (19-21), and has shown that LFIA sensitivities vary(20, 24). Importantly, in a large scale comparative study of the accuracy of four LFIAs, we observed a trade-off between sensitivity and specificity, with two devices being more sensitive and two more specific(20). All four devices studied in this work were more likely to give positive results for samples with high, rather than low, levels of antibodies following natural infection, and more specific devices detected low levels of antibodies less often than less specific devices (20).

The potential of lateral flow devices to predict individual risk of SARS-CoV-2 infection has not been evaluated. In this study our objective was to quantify directly the association of the results of four lateral flow antibody tests (Rapid Test Consortium’s AbC-19™ Rapid Test, OrientGene COVID IgG/IgM Rapid Test Cassette, SureScreen COVID-19 Rapid Test Cassette, and Biomerica COVID-19 IgG/IgM Rapid Test), and two laboratory-based immunoassays (Roche Elecsys® and EUROIMMUN Anti-SARS-CoV-2 ELISA), with subsequent symptom-driven PCR test positivity. We did this in a cohort of 2,826 unvaccinated UK keyworkers recruited to the EDSAB-HOME study (20, 21, 25, 26), and who were followed up for 200 days.

## Methods

### Study participants

The study population consisted of key workers recruited to the Evaluating Detection of SARS-CoV-2 AntiBodies at HOME (EDSAB-HOME) prospective cohort study (ISRCTN56609224) (27). Full descriptions of the participant characteristics, sample size considerations, and recruitment methods have been described previously(19-22, 25). Participants were recruited through their workplace into three streams: (A) Police and Fire & Rescue service keyworkers, (B) Health care keyworkers, both recruited regardless of previous SARS-CoV-2 infection status, and (C) Healthcare workers purposely recruited due to a history of previous RT-PCR positivity. The cohort contained 2,847 participants: 1,147 from Police and Fire (Stream A); 1,546 health care workers (HCW) (Stream B); and 154 from the healthcare worker previously COVID-19-positive test group (HCW-PP) (Stream C). Participants were recruited from two non-healthcare worker sites (one police station in Hutton, Lancashire, and one fire and rescue centre in Euxton, Lancashire) and six NHS acute hospitals (located in Milton Keynes, Gloucester, Cheltenham, York, Scarborough, and Rotherham), from 1 June to 26 June 2020 (28). We removed 21 individuals with incomplete laboratory test results, resulting in a total of 2,826 participants included in analysis. Recruitment took place in England in June 2020, at which point blood samples from participants were collected.

### Study endpoints

The cohort was monitored for the development of microbiologically-confirmed SARS-CoV-2 infection for the 200 days up to 24 January 2021, by linkage to a national database of SARS-CoV-2 results. Follow-up was stopped at this point due to the introduction of SARS-CoV-2 vaccination in the UK from December 2020 onwards(28).

The endpoint was testing positive for SARS-CoV-2 using nasal/throat PCR tests. In the period after recruitment, symptom-driven nasal/throat RT-PCR testing was available through state and employer routes for those with cough, fever, or disordered taste/smell; all such tests, irrespective of result, were recorded in a national database. Asymptomatic and lateral flow-based testing was available at the time, with positive lateral flow tests triggering PCR confirmation. Positive PCR tests triggered phone contact from NHS Test and Trace, a contact tracing service, who collected details of illness.

We obtained details about symptoms and circumstances associated with the positive test from

i. an optional weekly symptom questionnaire sent to volunteers;
ii. a symptom questionnaire sent retrospectively to all individuals with positive tests;
iii. NHS Test and Trace data.

### Lateral flow immunoassays (LFIAs)

Blood samples collected at recruitment from participants were analysed in the laboratory using four different lateral flow devices: the UK Rapid Test Consortium’s AbC-19™ Rapid Test (AbC-19); OrientGene COVID IgG/IgM Rapid Test Cassette (OrientGene); SureScreen COVID-19 Rapid Test Cassette (SureScreen); and Biomerica COVID-19 IgG/IgM Rapid Test (Biomerica)(19, 20). Each device provides a qualitative positive or negative result. The AbC-19, OrientGene, and SureScreen devices detect anti-Spike protein antibodies while the Biomerica device detects anti-Nucleoprotein antibodies. The OrientGene, SureScreen, Biomerica devices contain separate bands to detect IgG and IgM antibodies, while AbC-19 detects IgG antibodies only. In our analyses, we classified test results from OrientGene, SureScreen or Biomerica as positive if the IgG band was positive, disregarding the IgM bands. The laboratory protocol for conducting the lateral flow and laboratory-based immunoasssays has been described elsewhere(20).

### Laboratory assays

Blood samples from participants were also analysed with two commercial laboratory immunoassays: Roche Elecsys® Anti-SARS-CoV-2 (nucleocapsid (N)) and EUROIMMUN Anti-SARS-CoV-2 ELISA (IgG) assays (Spike (S) protein S1 domain)(29, 30). Roche Elecsys results were dichotomised at the manufacturer recommended threshold of 1.0. For EUROIMMUN, an immunoassay index lower than 0.8 is defined by the manufacturer as negative, an index between 0.8 and 1.0 is considered borderline, and an index greater than 1.0 is positive. In order to dichotomise the results, in this study an immunoassay index greater than 0.8 for EUROIMMUN was defined as positive.

### Blinding

The individuals who conducted the laboratory or lateral flow immunoassays could not access information about the samples, or results on any other assay. Participants were informed of EUROIMMUN serological results approximately one month after visiting the clinic, with a warning that this was not indicative of protection from disease. Participants were not informed of their Roche or LFIA results.

### Statistical analyses

Statistical analyses were conducted in R version 1.3.1056. Participant follow-up was divided into three periods based on national data describing the different waves of infection in the population at that time (Figure 1). For each antibody test, we describe the number of participants who tested positive for SARS-CoV-2, and observed rate per 100 person years, in each of these three periods, stratified by baseline test result. Poisson regression modelling with Firth penalisation was used to estimate incidence rates and incidence rate ratios (IRRs), describing the rate of SARS-CoV-2 positivity in individuals who tested antibody-positive compared to those who tested antibody-negative for each test, and confidence intervals. Adjusted IRRs were computed, by modelling in addition baseline risk factors for SARS-CoV-2 acquisition, specifically sex, ethnicity (white or non-white), age (continuous), high-risk occupation (medical or nursing staff), geographic region, and time-updated regional SARS-CoV-2 infection rates (average weekly incidence in the NHS region of residence of the participant). Kaplan-Meier survival curves were also produced, to describe the development of SARS-CoV-2 positivity over time in those who were antibody-positive or antibody-negative according to each test.

**Figure 1:**
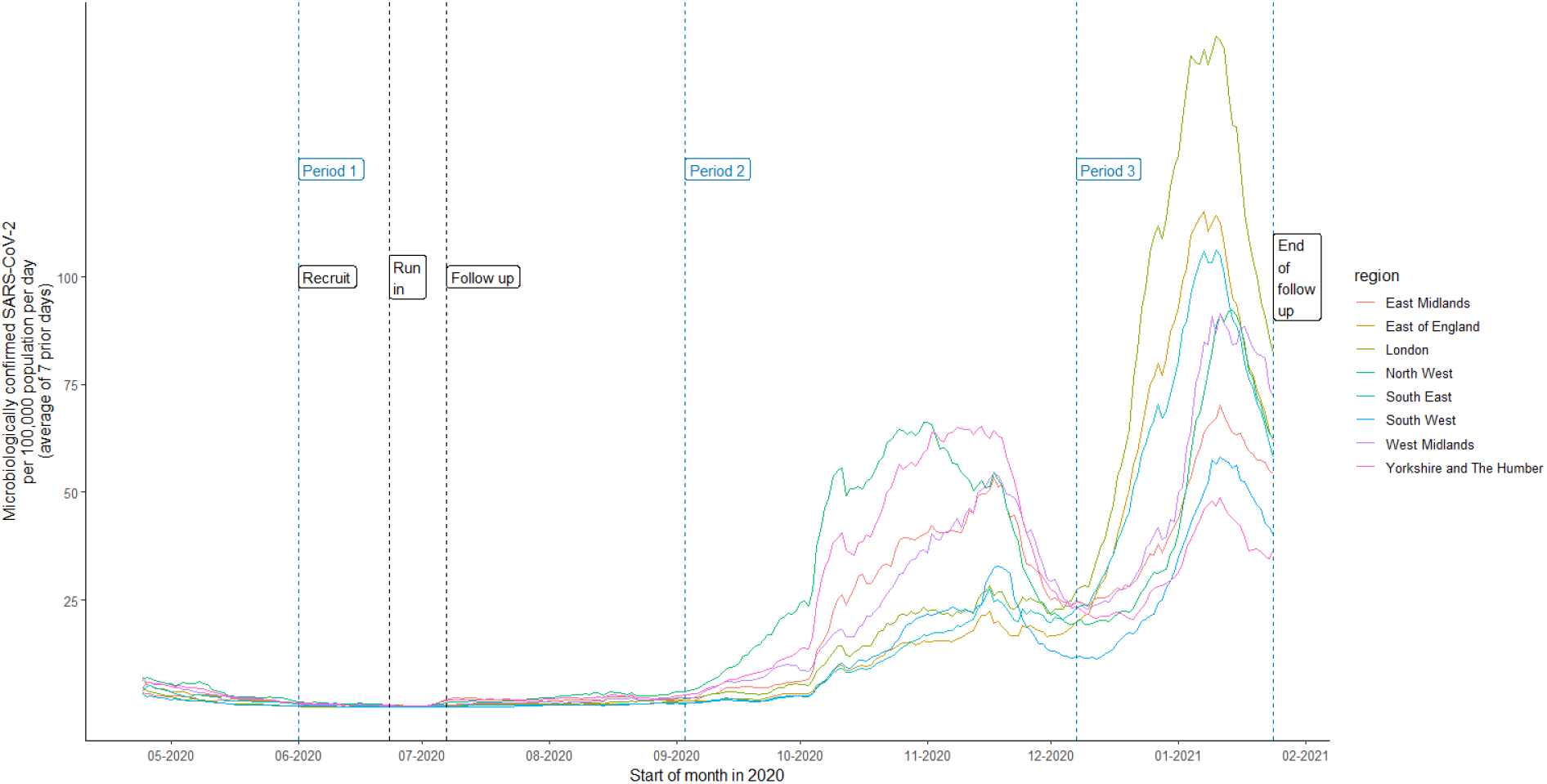
SARS-CoV-2 infections in England during the study period. National data showing the 7 day moving average of the case rate, per 100,000 people, stratified by region, throughout the study follow-up time. We split this into three periods based on the different waves of infection in the population: Period 1 = 01/06/2020 – 02/09/2020; Period 2 = 03/09/2020 – 06/12/2020; Period 3 = 07/12/2020 – 24/01/2021. Recruitment took place from 01/06/2020 – 30/06/2020. There was a run in period of 2 weeks after recruitment where any positive SARS-CoV-2 results were excluded to ignore individuals who were already infected. Follow-up was for 200 days post recruitment. Data on population size obtained from Office for National Statistics. Data on SARS-CoV-2 cases obtained from UK Government.

In additional exploratory analyses, the relationship between the thresholds used to define antibody positivity for the Roche Elecsys and EUROIMMUN laboratory-based immunoassays and the adjusted IRRs for PCR positivity was explored graphically.

### Ethics

The EDSAB-HOME study was approved by NHS Research Ethics Committee (Health Research Authority, IRAS 284980) on 02/06/2020 and PHE Research Ethics and Governance Group (REGG, NR0198) on 21/05/2020. All participants gave written informed consent.

## Results

### SARS-CoV-2 positivity

The study follow-up period ran until 24 January 2021. After a period with low levels of infection (period 1), two waves of intense SARS-CoV-2 transmission in England occurred (periods 2,3) (Figure 1)(31). During follow-up, 285/2,826 (10%) of participants had positive SARS-CoV-2 RT-PCR tests (Table 1). Symptom data were available on 216, of whom 15 (7%) reported having no symptoms in questionnaires when they were tested.

**Table 1:** Numbers of SARS-CoV-2 PCR tests positive by antibody status. Number of SARS-CoV-2 RT-PCR positives and observed rate of PCR positivity amongst participants who tested antibody-negative and participants who tested antibody-positive with each LFIA and laboratory immunoassay (n = 2826). *Adjusted for sex, ethnicity (white or non-white), age (continuous), high-risk occupation (medical or nursing staff), region, average weekly SARS-CoV-2 infection incidence rate in NHS region of residence of participant. □ Crude observed rate before application of Firth penalisation of 0.0. CI: Confidence interval.

|  |  | **AbC-19** | **SureScreen** | **OrientGene** | **Biomerica** | **Roche** | **EUROIMMUN** |
| --- | --- | --- | --- | --- | --- | --- | --- |
| **Antibody test negatives** | **Number testing antibody negative** | **2261** | **2269** | **2203** | **2186** | **2219** | **2237** |
|  | Number of PCR positives in Period 1 | 3 | 3 | 3 | 3 | 3 | 3 |
|  | Number of PCR positives in Period 2 | 109 | 109 | 107 | 103 | 108 | 108 |
|  | Number of PCR positives in Period 3 | 173 | 171 | 169 | 170 | 172 | 171 |
|  | Total number of PCR positives | 285 | 283 | 279 | 276 | 283 | 282 |
|  | Person years at risk | 1456.7 | 1462.0 | 1419.3 | 1409.0 | 1429.4 | 1441.1 |
|  | **Observed rate of PCR positivity per 100 person years with Firth penalisation (95% CI)** | **19.6 (17.5–22.0)** | **19.4 (17.3–21.8)** | **19.7 (17.5–22.1)** | **19.6 (17.4–22.1)** | **19.8 (17.7–22.3)** | **19.6 (17.4–22.0)** |
| **Antibody test positives** | **Number testing antibody positive** | **565** | **557** | **623** | **640** | **607** | **589** |
|  | Number of PCR positives in Period 1 | 0 | 0 | 0 | 0 | 0 | 0 |
|  | Number of PCR positives in Period 2 | 0 | 0 | 2 | 6 | 1 | 1 |
|  | Number of PCR positives in Period 3 | 0 | 2 | 4 | 3 | 1 | 2 |
|  | Total number of PCR positives | 0 | 2 | 6 | 9 | 2 | 3 |
|  | Person years at risk | 370.1 | 364.7 | 407.5 | 417.8 | 397.3 | 385.6 |
|  | **Observed rate of PCR positivity per 100 person years with Firth penalisation (95% CI)** | **0.1 □ (0.0–2.2)** | **0.7 (0.2–2.4)** | **1.6 (0.7–3.4)** | **2.3 (1.2–4.3)** | **0.6 (0.2–2.2)** | **0.9 (0.3–2.6)** |
| **Unadjusted rate ratio with Firth penalisation (95% CI)** |  | 0.00 (0.00–0.02) | 0.03 (0.02–0.05) | 0.08 (0.06–0.11) | 0.10 (0.07–0.13) | 0.03 (0.02–0.05) | 0.05 (0.03–0.07) |
| **Adjusted \* rate ratio with Firth penalisation (95% CI)** |  | 0.00 (0.00–0.01) | 0.03 (0.02–0.05) | 0.07 (0.05–0.10) | 0.09 (0.07–0.12) | 0.02 (0.01–0.04) | 0.04 (0.03–0.06) |

Overall, 3/285 (1%) PCR positives were reported in period 1, 109/285 (38%) in period 2, and 173/285 (61%) in period 3. Kaplan Meier survival curves for each LFIA are shown in Figure 2. The crude rate of PCR positivity amongst antibody-positive participants was lowest for AbC-19 (0 observed events, estimated rate with Firth penalisation of 0.1 per 100 person-years (95%CI: 0.0–2.2)), followed by Roche (0.6 (95%CI: 0.2–2.2)), SureScreen (0.7 (95%CI: 0.2–2.4)), EUROIMMUN (0.9 (95%CI: 0.3–2.6)), OrientGene (1.6 (95%CI: 0.7– 3.4)), and Biomerica (2.3 (95% CI: 1.2-4.3)).

**Figure 2:**
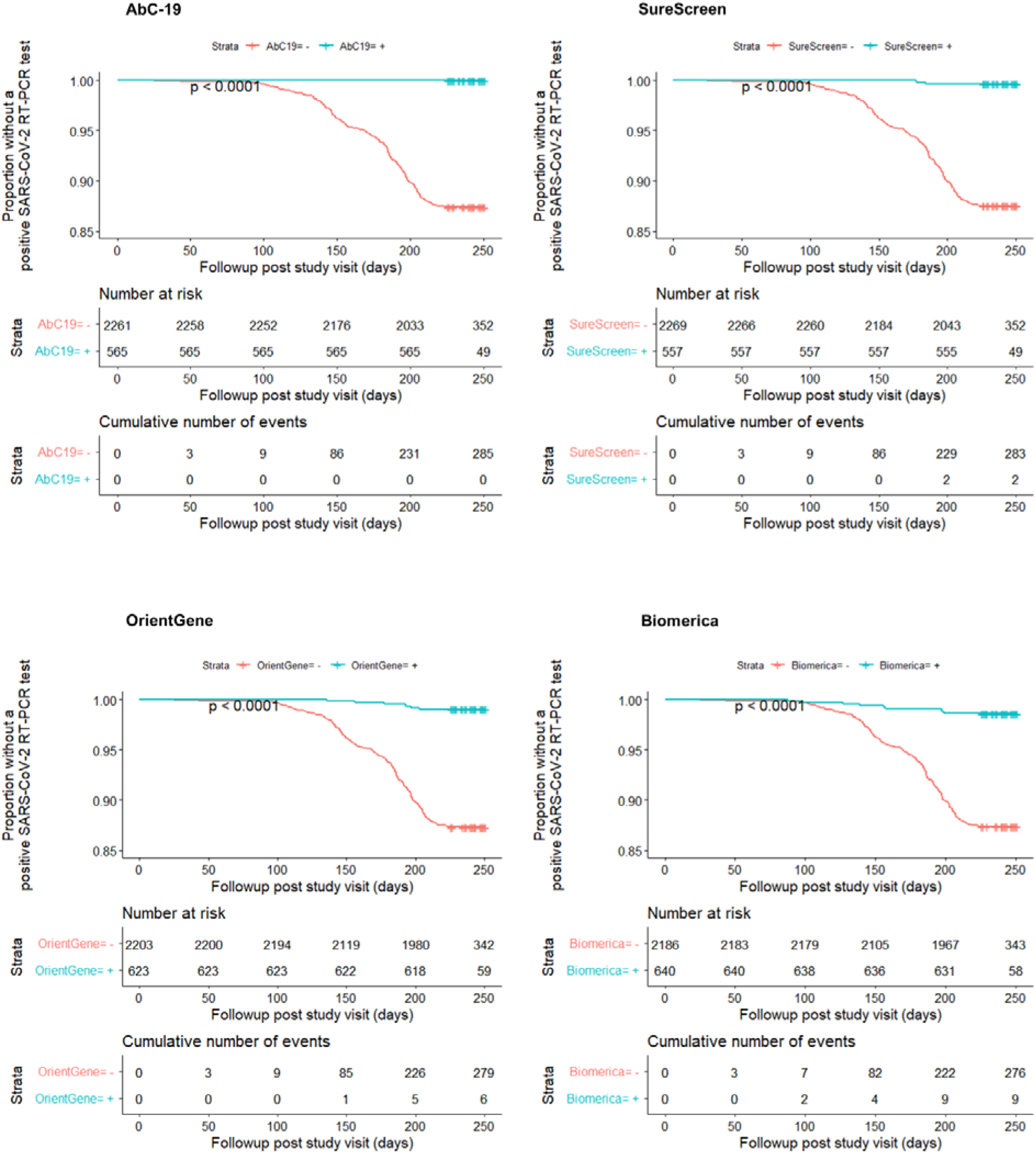
Time to positive SARS-CoV-2 RT-PCR test stratified by antibody test result for each LFIA. Kaplan Meier survival curves showing the proportion of participants without a positive SARS-CoV-2 RT-PCR test throughout the follow up period, stratified by antibody test result for each lateral flow immunoassay.

### Association of results and subsequent SARS-CoV-2 infection

The adjusted rate ratio for PCR positivity was lowest for the AbC-19 device (0.00 (95%CI: 0.00–0.01)), followed by Roche (0.02 (95%CI: 0.01–0.04)), SureScreen (0.03 (95%CI: 0.02– 0.05)), EUROIMMUN (0.04 (95%CI: 0.03–0.06), OrientGene (0.07 (95%CI: 0.05–0.10)) and Biomerica (0.09 (95%CI: 0.07–0.12)), but with overlapping CIs. (Table 1, Figure 3).

**Figure 3:**
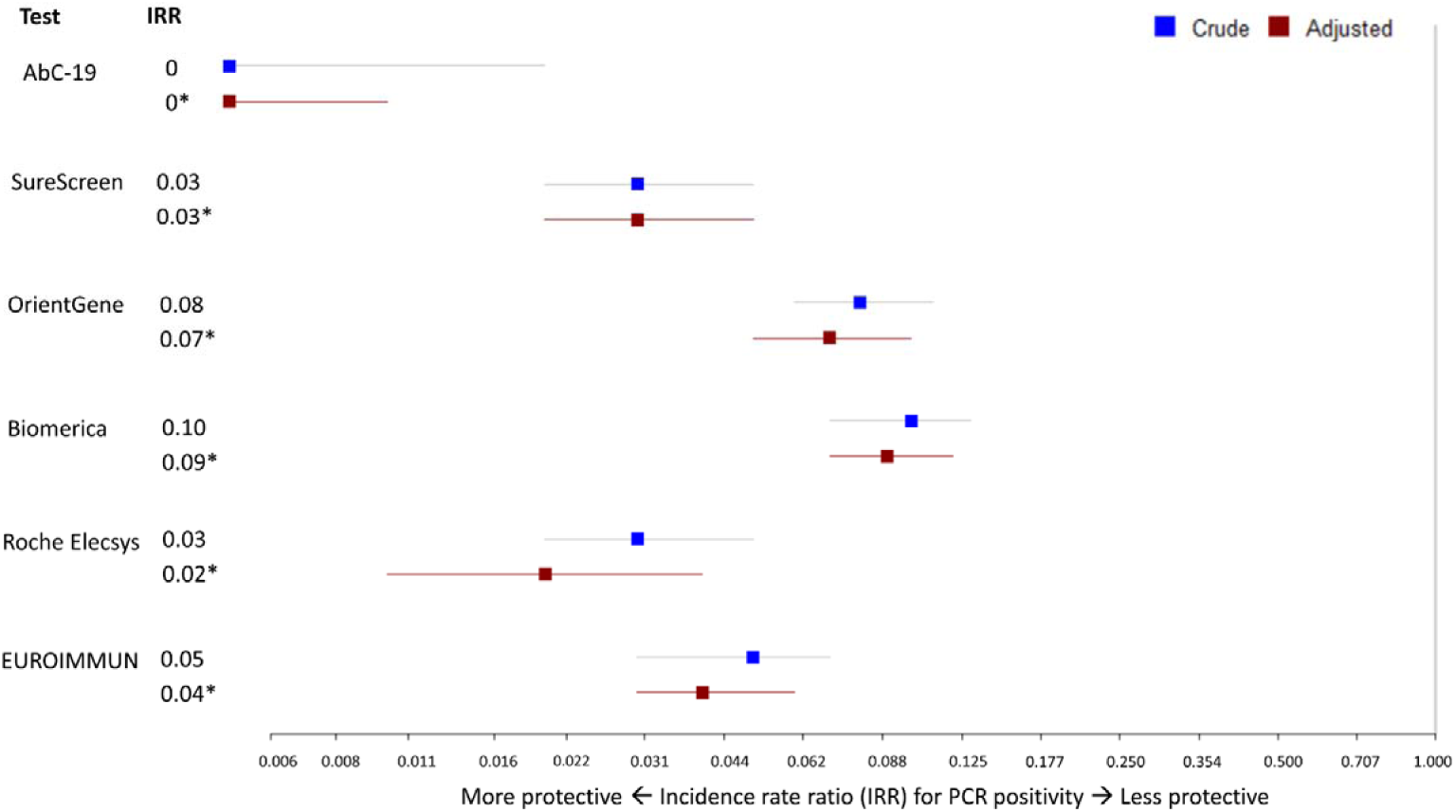
Incidence rate ratios for SARS-CoV-2 PCR positivity by test result. Forest plot showing observed and adjusted (*) incidence rate ratios (IRRs, aIRRs) (95% confidence intervals) for PCR positivity in those who tested antibody-positive in each test compared to those who tested antibody-negative. Adjustment included sex, ethnicity (white or non-white), age (continuous), high-risk occupation (medical or nursing staff), geographic region, and average weekly SARS-CoV-2 infection incidence rate in the NHS region of residence of the participant. Roche Elecsys results of an immunoassay signal greater than 1.0 were considered positive. EUROIMMUN results of an immunoassay index greater than 0.8 were considered positive.

### Antibody positivity thresholds

Since the sensitivity of lateral flow devices for the detection of low levels of SARS-CoV-2 antibodies is known to differ between devices, we explored how the association varied when alternative test positivity thresholds were used for the laboratory immunoassays (20). In Figure 4, we present the adjusted incidence rate ratios for both Roche and EUROIMMUN laboratory-based immunoassays, dichotomised at various different assay signals. The strength of association between test positivity and subsequent infection was somewhat stronger when higher positivity thresholds were used (Figure 4).

**Figure 4:**
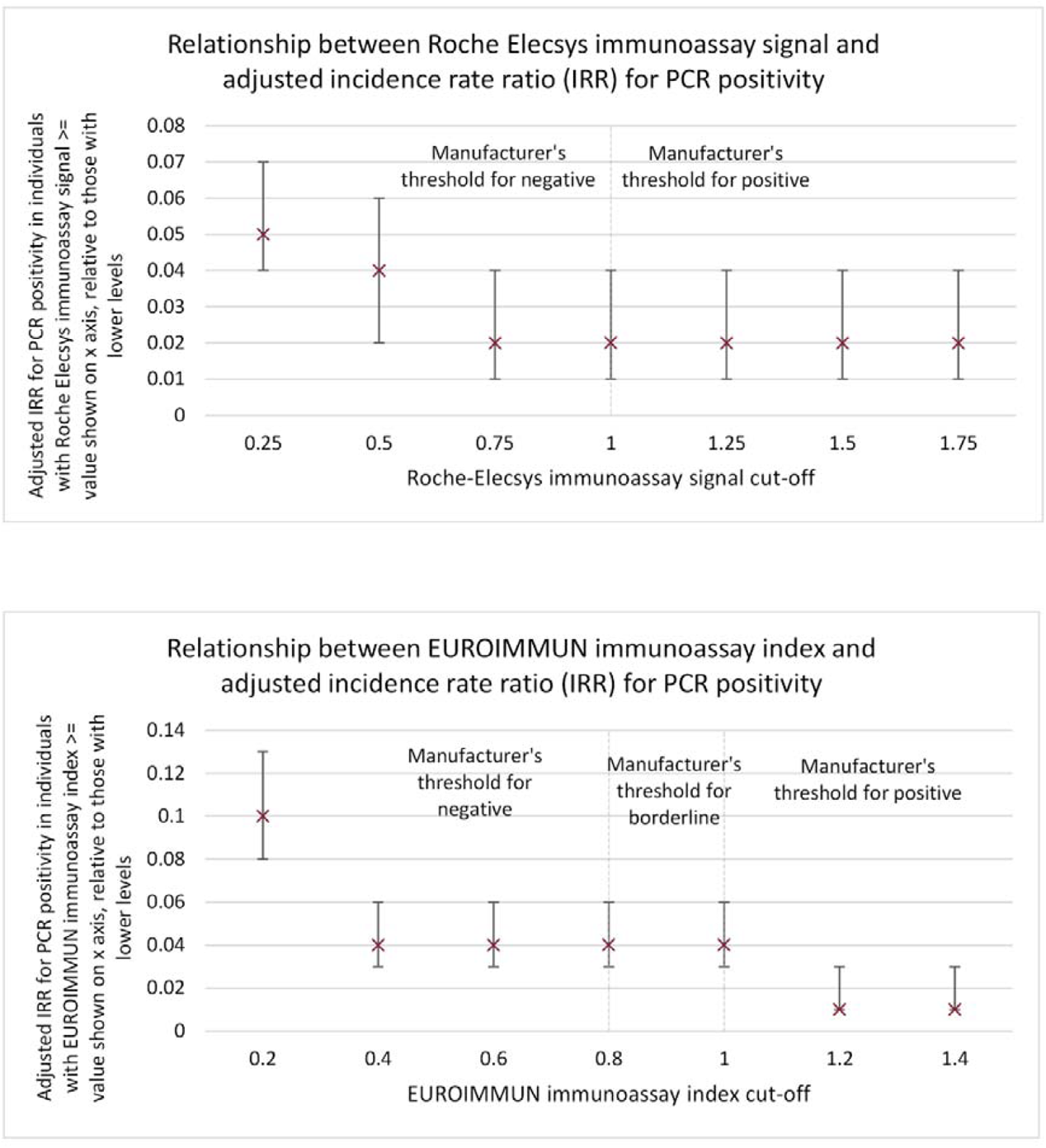
Roche Elecsys and EUROIMMUN results and PCR positivity. Adjusted incidence rate ratios (IRRs) for PCR positivity when Roche Elecsys (upper panel) and EUROIMMUN (lower panel) assays, measuring anti-Nucleoprotein and anti-Spike S1 antibody reactivity respectively, are dichotomised at a series of different thresholds. For example, the incidence rate ratio shown on the x-axis at 1 refers to the incidence rate ratio in individuals with a baseline antibody level of 1 or higher, relative to those below. Adjustment included sex, ethnicity (white or non-white), age (continuous), high-risk occupation (medical or nursing staff), geographic region, and average weekly incidence rate in the NHS region of residence of the participant. Error bars show 95% confidence intervals.

## Discussion

This study demonstrates that participants who tested positive for SARS-CoV-2 antibodies with any of the four lateral flow immunoassays (AbC-19, SureScreen, OrientGene, and Biomerica) or two laboratory immunoassays (Roche and EUROIMMUN) detecting anti-SARS-CoV-2 antibodies had a lower rate of subsequent SARS-CoV-2 infection than individuals who did had no detectable antibodies. Adjusted incidence rate ratio (95% CI) estimates for disease were 0.00 (0.00,0.01) and 0.03 (0.02, 0.05) for the two most predictive lateral flow devices.

While this study shows a strong association between lateral flow device results and subsequent SARS-CoV-2 infection, it has several limitations. Firstly, it applies to a historical cohort of unvaccinated individuals. Secondly, lateral flow devices were applied and read using stored plasma, obtained by venesection, in a laboratory setting by trained professionals, rather than relying on measurement in the field by the public using finger-prick technology. This arrangement had the advantage that the subjects did not know their lateral flow device results, minimising potential bias, but leaves open the possibility that performance on finger prick samples might differ from that in the laboratory setting we used. This concern has been addressed for the SureScreen device,, for which data has now been published showing very similar accuracy of the SureScreen device on finger-prick samples taken from individuals and serum samples analysed in a laboratory (24). Finally, some healthcare workers in the cohort received a single vaccine dose in late December 2020 and early January 2021 in the UK, in the final weeks of our follow-up period; it is possible that some individuals acquired protection through vaccination in the last weeks of follow-up, so the true protection associated with LFIA positive tests may be higher than we observed.

Lateral flow devices have very different reported accuracy. This is likely to be explicable in part by device design, and in part by selection of samples used for device evaluation(20, 32, 33). Of the four LFIAs included in our study, comparative testing on a large, well-characterised sample set showed Abc-19 and SureScreen were more specific, whilst Biomerica and OrientGene were more sensitive and detected lower levels of SARS-CoV-2 antibodies, including those at which disease risk is elevated (Figure 4)(20). Therefore, the differential sensitivities of these devices may explain the variation in disease risk associated with testing LFIA-positive using various different lateral flow devices. This observation may inform device choice and design decisions when lateral flow devices are optimised: during development, specificity/sensitivity trade-offs operate. Devices that are less sensitive but more specific, and which perform well in useability tests (24), may be more useful for predicting disease protection.

Overall, these data suggest that the more specific LFIA devices used here may have a role in surveillance programs assessing population protection in unvaccinated individuals, informing the debate about the risk to populations, and perhaps in individual risk assessment(4, 5). As part of such work, an ongoing programme of field studies would be required to show whether the LFIA-associated protection seen in this study extends to self-read and healthcare worker-read tests, to currently prevalent SARS-CoV-2 strains which may differ from the strains circulating during this study (34, 35), and (if surveillance of vaccinated individuals were contemplated) to vaccinated individuals.

## Data Availability

The data analysed is available from the corresponding author on request.

## Author contributions

Conceptualisation: HEJ, STP, RM, IO, DW; Data curation: Amy Trindall (AT1), DW, RM; Formal Analysis; LF, Adam Trickey (AT2),DW; Funding: IO; Investigation: AT2,RM,DW; Methodology: AT2, HEJ, STP, DW; Resources: PM, JB, AH, NT, IR; Writing first draft: LF, DW; EDSAB-HOME clinical investigators (PM, JB, AH, NT, IR) contributed to study design and resources (supporting access to volunteers). Reviewed and revised manuscript: all authors. Access to underlying data: RM, DW, LF.

EDSAB-HOME investigators: Philippa Moore and John Boyes (Gloucestershire Hospitals NHS Foundation Trust), Anil Hormis (The Rotherham NHS Foundation Trust), Neil Todd (York Teaching Hospital NHS Foundation Trust), and Ian Reckless (Milton Keynes University Hospital NHS Foundation Trust).

## Data availability

The data analysed is available from the corresponding author on request.

